# The association of a streamlined and updated stroke protocol with improved secondary prevention and outcome

**DOI:** 10.1101/2023.05.09.23289745

**Authors:** Almond Leung, Richard J Fong, Karim M Mahawish

## Abstract

**Background:** Stroke recurrence rates following an index event remain high compared with the baseline population. Evidence-based stroke treatments reduce this risk.

**Aims:** To determine the effect of an updated and streamlined hospital stroke guideline on prescribing practices and stroke recurrence rates at 3 months.

**Methods:** Hospital registries were searched for ICD-10 codes for transient ischaemic attack (TIA) and ischaemic stroke between July 2019 and July 2020. Data on patient demographics, discharge documentation, and other outcomes of interest were dichotomised into pre and post-intervention.

**Results:** 396 patients were identified. There was a significant reduction in the stroke recurrence rate at 3 months post guideline update (8.3% vs 2.2%, OR 0.24, p<0.01). There were significant improvements in prescriptions for statins (66.5% vs 81.2%, p≤0.01) and antihypertensives (40.7% vs 50.7%, p=0.05), and documentation of lipid and blood pressure targets in discharge letters. There was a trend towards greater use of dual antiplatelet therapy (25.2% vs 34.1%, p=0.057).

**Conclusion:** In this audit, we found an association between the guideline update and improved patient outcomes and prescribing practices. We were unable to directly attribute the reduction in stroke recurrence to any single factor. This may be a chance finding and warrants investigation in other settings.

## Introduction

Stroke is a leading cause of death and disability in New Zealand. It is estimated that approximately 11,000 people experienced a stroke in 2020.^1^ A 40% rise in stroke incidence is anticipated over the next decade.^1^

The pace of discovery of new evidence for effective stroke treatments has led to the development and adoption of the online ‘living’ Australian and New Zealand clinical guidelines for stroke management.^2^ It is unclear whether health care staff are more likely to refer to national or local guidelines to guide management decisions.

Despite advances in stroke management, recurrence rates are high following an index TIA or stroke. Secondary stroke prevention is essential in mitigating this risk. A population-based cohort study over a 20-year period demonstrated a reduction in five-year stroke recurrence rates from 18% to 12% as a direct result of improvements in stroke secondary prevention.^3^

Our Institution is a secondary provincial hospital in New Zealand serving a population of 186,190.^4^ It has a stroke service led by a stroke physician and consists of a clinical nurse specialist, junior medical staff, and members of the multidisciplinary team. Our institution’s stroke management guidelines were last published in May 2018, making some aspects obsolete; an update was necessary to reflect modern practice. The amalgamation of the two existing guidelines (initial assessment and admission guideline and inpatient management guideline) also aimed to simplify the documents for conciseness and readability. The changes made are summarised in table 1. There is a paucity of data regarding stroke recurrence rates and the appropriateness of secondary stroke prevention prescribing at our institution.

**Table 1:** Summary of changes made to the guideline

| New recommendations | Amendments | Deletions |
| --- | --- | --- |
| Clarity on timing of CT head (indications for immediate CT head listed) in body of guideline rather than appendix. | Amalgamation of two existing guidelines into one document (Clinical guideline for stroke/ TIA management) | Recommendation for aspirin and dipyridamole as an alternative to clopidogrel |
| Consideration of early referral for patients at risk of malignant middle cerebral artery syndrome | Addition of quick dial number for Capital and Coast District Health Board | Recommendation for simvastatin |
| Dual antiplatelet therapy for high-risk TIA and minor stroke | Intravenous anti-hypertensive options broadened | Avoidance of anti-embolic stockings post stroke |
| Post stroke blood glucose level targets | Reduced word count from 3236 to 2240 and improved readability scores |  |
| Systolic blood pressure and low density lipoprotein targets |  |  |

We performed this audit to determine the effect of the guideline update on prescribing practices and discharge documentation of blood pressure and lipid treatment targets. Further, we investigated stroke recurrence and bleeding complications in the pre and post-intervention periods in the three months following the index event. Stroke recurrence was defined as any recurrent stroke occurring more than 24 hours after the incident stroke, irrespective of vascular territory. This definition was recommended by Coull et al^5^ to avoid the underestimation of stroke recurrence and to allow comparison amongst studies. The Trial of Org 10172 in acute ischaemic stroke (TOAST) classification was used to determine stroke subtype.^6^ The Bleeding Academic Research Consortium (BARC) types 2, 3, and 5 were used to define major haemorrhage.^7^ These categories include instances of haemorrhage which require medical attention or are fatal. The audit standards were based on the updated hospital TIA and stroke guideline, which were taken from the Australian Clinical Guidelines for Stroke Management.^8^

## Methods

Hospital registries were searched for ICD-10 codes G45.9 (transient ischaemic attack, unspecified) and I63.0, I63.1, I63.3, I63.4, I63.9 (cerebral infarction due to thrombosis or embolism of the precerebral and cerebral arteries, and cerebral infarction, unspecified) to identify the National Health Index details of patients. Patients aged 16 years or older presenting to our institution’s Emergency Department or admitted to hospital with a clinical diagnosis of TIA or ischaemic stroke from 22/7/19 – 22/1/20 (pre-intervention) and 23/1/20 – 23/7/20 (post-intervention) were eligible for inclusion. Patients with haemorrhagic stroke, non-stroke presentations, death during the index admission, or those discharged for end-of-life care were excluded. Electronic patient records were used for data collection and paper files were requested if required. Documentation from emergency department visits and hospital discharge summaries in the 90 days following the index admission were reviewed to determine the primary efficacy outcome of stroke recurrence and the primary safety outcome of major bleeding.

Data on demographics, admission date, presence of atrial fibrillation, secondary prevention prescribing, stroke physician input, stroke recurrence, and bleeding complications were recorded. If the management of a patient deviated from the guideline but the rationale was documented and appropriate, it would be considered in accordance with the standard, e.g. dual antiplatelet treatment for three months in patients with symptomatic intracranial stenosis.

Continuous and categorical variables are expressed as means and frequencies respectively. Baseline characteristics were compared between the pre and post-intervention groups; categorical variables were compared using Fisher’s exact or χ^2^ test as appropriate. Univariate and multivariate analyses and logistic regression were performed to test for significant associations. A p-value of ≤0.05 was considered statistically significant. R was used to perform statistical analysis. This audit was exempt from ethics approval following institutional review. The SQUIRE checklist was used when writing our report.^9^

## Results

167 TIA and ischaemic stroke presentations (pre-intervention) and 229 presentations (post-intervention) were eligible for inclusion. Patient demographics and TOAST classification are detailed in table 2.

**Table 2.** Patient demographics and TOAST classification

|  | PRE-INTERVENTION<br>N (%) | POST-INTERVENTION<br>N (%) | *P* VALUE |
| --- | --- | --- | --- |
| Mean age (range) years | 73.8 (21 – 99) | 73.7 (28 – 99) | 0.63 |
| Ethnicity |  |  | 0.76 |
| New Zealand European | 143 (85.6%) | 193 (84.3%) |  |
| Maori | 17 (10%) | 22 (9.6%) |  |
| Asian | 2 (1.2%) | 7 (3%) |  |
| Pacific Islander | 2 (1.2%) | 4 (1.7%) |  |
| Other | 3 (1.8%) | 3 (1.3%) |  |
| Male gender | 82 (49.1%) | 104 (45.4%) | 0.48 |
| Atrial fibrillation | 55 (32.9%) | 55 (24%) | 0.05 |
| TOAST classification |  |  | 0.61 |
| Small vessel disease | 47 (28.1%) | 52 (22.7%) |  |
| Large vessel disease | 15 (9%) | 28 (12.2%) |  |
| Cardioembolism | 32 (19.2%) | 39 (17%) |  |
| Undetermined | 46 (27.5%) | 48 (21%) |  |
| Other mechanism | 2 (1.2%) | 5 (2.2%) |  |
| TIA | 25 (15%) | 57 (24.9%) |  |

Following the guideline update, there was a significant reduction in stroke recurrence rates at 3 months (8.3% vs 2.2%, OR 0.24 [95%C.I. 0.07-0.73], p<0.01) (Table 3). A greater proportion of patients in the post-intervention group were prescribed statin therapy in accordance with the guideline (66.5% vs 81.2%, p≤0.01) and had more changes to antihypertensive medication (40.7% vs 50.7%, OR 1.5 [95%CI 1-2.24], p=0.05). There was a trend toward more frequent use of dual antiplatelet medication (aspirin and clopidogrel) in the post-intervention group (25.2% vs 34.1%, OR 1.54 [95%CI 0.99–2.39], p=0.057). A significantly higher proportion of patients had blood pressure and lipid targets documented in their discharge summary in the post-intervention group (6.6% vs 16.6%, p≤0.01) and (4.2% vs 14.8%, p≤0.01) respectively. In both groups, anticoagulation was prescribed for all patients with an indication.

**Table 3.**
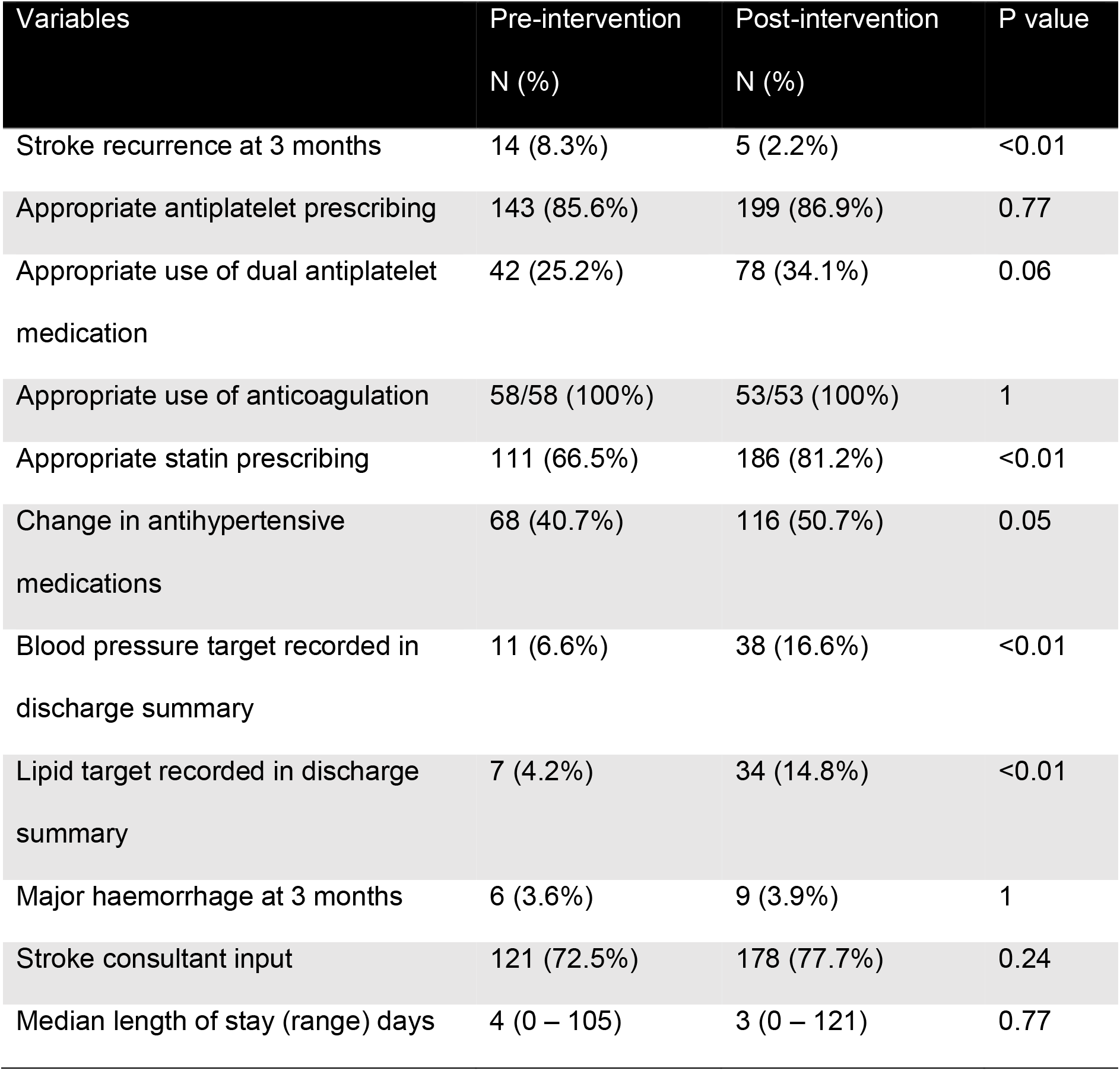
Summary of clinical outcomes

| Variables | Pre-intervention<br>N (%) | Post-intervention<br>N (%) | P value |
| --- | --- | --- | --- |
| Stroke recurrence at 3 months | 14 (8.3%) | 5 (2.2%) | <0.01 |
| Appropriate antiplatelet prescribing | 143 (85.6%) | 199 (86.9%) | 0.77 |
| Appropriate use of dual antiplatelet medication | 42 (25.2%) | 78 (34.1%) | 0.06 |
| Appropriate use of anticoagulation | 58/58 (100%) | 53/53 (100%) | 1 |
| Appropriate statin prescribing | 111 (66.5%) | 186 (81.2%) | <0.01 |
| Change in antihypertensive medications | 68 (40.7%) | 116 (50.7%) | 0.05 |
| Blood pressure target recorded in discharge summary | 11 (6.6%) | 38 (16.6%) | <0.01 |
| Lipid target recorded in discharge summary | 7 (4.2%) | 34 (14.8%) | <0.01 |
| Major haemorrhage at 3 months | 6 (3.6%) | 9 (3.9%) | 1 |
| Stroke consultant input | 121 (72.5%) | 178 (77.7%) | 0.24 |
| Median length of stay (range) days | 4 (0 – 105) | 3 (0 – 121) | 0.77 |

With respect to safety endpoints, there was no difference in the rates of major haemorrhage between the two groups (3.6% vs 3.9%, p=1). Univariate analysis of each of the predictors used in the full logistic regression model failed to identify any one factor which led to our statistically significant results. Further, multivariate logistic regression analysis (Table 4) failed to identify any associations with the reduction in stroke recurrence (all p-values >0.05). Finally, we were unable to identify associations between TOAST classification, length of stay, or age on adjusted analyses (Table 4). McFadden’s pseudo-R^2^ was 0.017 (non-significant).

**Table 4.** Summary of logistic regression testing

| Variables | P value |
| --- | --- |
| Antiplatelet on discharge | 0.88 |
| Dual antiplatelet therapy use | 0.85 |
| Lipid lowering medication | 0.35 |
| Change in antihypertensive medication | 0.77 |
| Blood pressure target on discharge summary | 0.48 |
| Lipid target on discharge summary | 0.91 |
| Stroke consultant input | 0.85 |
| TOAST: |  |
| Large vessel atherosclerosis | 0.91 |
| Small vessel occlusion | 0.95 |
| Stroke of other determined aetiology | 0.28 |
| Stroke of undetermined aetiology | 0.98 |
| TIA | 0.61 |
| Length of stay | 0.35 |
| Age | 0.39 |

## Limitations

This audit has several limitations. Full data on comorbidities was not collected on each patient and may serve as a confounder, e.g. due to the retrospective nature of this audit, the National Institutes of Health Stroke Scale could not be obtained in the majority of patients.

Further, we have no data on adherence to medications or follow-up monitoring on discharge to ensure the pre-specified lipid and blood pressure targets were achieved. Other potential unmeasured confounders include concurrent public awareness campaigns or changes to hospital services during the audit period. Data obtained was limited to hospital registries; patients consulting other health providers with TIA or stroke presentations, loss to follow-up, and competing risks were not accounted for. This was a single centre audit at a secondary provincial hospital setting so the results may not be generalisable to other sites. Finally, this audit was carried out immediately after the introduction of the updated guideline so awareness of the guideline was heightened.

## Discussion

This audit demonstrated a four-fold reduction in the risk of stroke recurrence associated with the guideline update. Other significant findings included an improvement in statin and antihypertensive prescribing and the inclusion of blood pressure and lipid targets in the discharge summary. The rates of major haemorrhage also did not differ significantly.

McFadden’s pseudo-R^2^ was 0.017 indicating that about 1.7% of the variation in the data is explained by the logistic regression model. This is consistent with the non-significance of all the predictors: Logistic regression analysis failed to identify any single factor which was associated with these findings. This may be due to the low number of stroke recurrences observed over the audit period, limiting further statistical analysis. While our findings may be due to chance, they may be the effect of the entire care bundle rather than one individual factor.

The increased use of statins post-intervention is noteworthy. It is unlikely to be causal in the reduction of stroke recurrence rates; In the randomized controlled trial of early (within 24 hours) versus delayed (within seven days) statin therapy in patients with acute ischemic stroke (ASSORT Trial), there was no significant reduction in stroke recurrence at 90 days.^10^ In this study, moderate-intensity statins were used (i.e. atorvastatin 20mg OD, rosuvastatin 5mg OD, etc.). Similarly, in the stroke prevention by aggressive reduction in cholesterol levels (SPARCL), trial, no benefit was seen in the first year of treatment. In this trial however, treatment was started on average three months after the index event.^11^ There is observational data that early statin use in acute ischaemic stroke may be associated with significant reductions in stroke recurrence.^12^ Since therapeutic benefits in the management of acute stroke and TIA are front-loaded, the benefit of early high-intensity statin after acute ischaemic stroke warrants further exploration.

The risk of a recurrent ischaemic stroke after the initial presentation with TIA or stroke is highest in the first few months. In a recently published population-wide cohort study of adults with first stroke hospitalisation in Australia and New Zealand, the cumulative incidence of stroke recurrence was 7.8% at three months.^13^ Internationally, the acute rate varies between studies due to heterogeneity in methodology and definition of stroke recurrence and ranges from 3-15%.^14-16^ The proportion of patients at our institution discharged on statins, anti-hypertensives and antithrombotic agents are comparable to stroke centres in Australia.^17^ All eligible patients at our institution were prescribed anticoagulation if indicated, while in Australian stroke centres 74% of patients were prescribed anticoagulants for atrial fibrillation.^17^ Interestingly, in the ‘Effect of urgent treatment of transient ischaemic attack and minor stroke on early recurrent stroke (EXPRESS study)’, the primary outcome of 90-day risk of recurrent stroke reduced from 10.3% to 2.1%,^18^ similar findings to our study and suggesting more effective secondary prevention post-intervention.

The guideline update amalgamated two pre-existing guidelines, leading to a reduction in word count from 3236 to 2240 and an improvement in readability score as calculated as the Flesch Reading Ease Score (FRES). This score takes into account sentence and word length, with higher scores indicating easier readability.^19^ The prior guidelines had FRES of 18.19 and 35.53, while the updated guideline had a FRES of 29.05.

## Conclusion

In this audit, we found a statistically significant reduction in stroke recurrence after the TIA and stroke management guideline update at our institution. While other temporal factors and confounders likely played a role, the positive association of the guideline update and reduced stroke recurrence suggests that frequent guideline updates if for no other reason than raising awareness are important. The next step would be to conduct a repeat audit to assess for ongoing adherence to the guideline and time trends in stroke recurrence rates. Approximately 4% of patients in the cohort experienced major bleeding within three months of the index event. A further area of study would be to determine risk factors associated with a higher risk of haemorrhage. This may allow safer, more individualised secondary prevention prescribing.

## Implications

There is an association between updating and simplifying local clinical guidelines and improved prescribing practices and patient outcomes.

This study suggests that hospital guidelines should be updated to reflect modern evidence-based practice and raise awareness.

## Data Availability

All data produced in the present study are available upon reasonable request to the authors

## Acknowledgements

Nil

## Statement of contribution

AL conceived the project, collected data and drafted the manuscript. RJF performed the statistical analysis and reviewed the manuscript. KMM assisted with statistical analysis and drafting the final manuscript.

